# Efficacy and Safety of Magnetic Seed Pre-Operative Non-Palpable Breast Lesion Localization: a series of 37 consecutive cases

**DOI:** 10.1101/2023.08.11.23292939

**Authors:** M. Bouvier, Y Abrahami, I. Belaroussi, MR El Bejjani, S. Béranger, M. Cascarino, R. Afriat, N Lotersztajn, S. Alran

## Abstract

**Context:** 40% of breast surgery patients have a lesion that requires preoperative localization, a process that demands close cooperation between radiological and surgical team. Magnetic seed localization is a new tracking technique which does not require programming the day before or on the day of the intervention. The aim of our study was to evaluate the efficacy and safety of magnetic localization of non-palpable breast lesions.

**Methods and patients:** This is a study of 39 consecutive preoperative ultrasound-guided implantations of a magnetic seed (MS) in 37 patients, for non-palpable breast lesions, performed at the Breast Center at Saint-Joseph Hospital in Paris, France, between May 15^th^ and December 21^st^, 2018. One patient who was operated on for papillomatous lesions had a double magnetic seed implanted. In the operating room, the MS was percutaneously localized by a magnetic probe. The ex-vivo magnetism was noted and the removed tissue was sent to radiology to look for the MS, after which it was sent for histopathological examination. All localized lesions had previously been biopsied, and there were 29 infiltrative cancers, 7 atypical lesions, and 3 benign lesions. The sentinel node was identified by super paramagnetic iron peroxide in 11 cases, and by isotopes in the 18 others.

**Results:** Our patients were on average 57 years old (33-86 years old). All magnetic localization was realized using ultrasound. The mean ultrasound size of the lesions was 12.7 mm (5-34mm). The period of time from implantation to surgery varied from 0 to 21 days. The localization method was characterized by a rapid pose, facilitated by the excellent luminosity of the needle for the tracking. No compression pad was needed, optimizing the implementation and quality of the control mammography. The mean time for the tissue resection from incision to excision was 15 minutes for the first 10 cases. On the radiography of removed tissue: the clip was present in 38 out of 39 cases. One failure was registered, in relation to loss of the clip, found in the tumorectomy limits, in the patient with the double localization procedure. However, the target was effectively removed and detected histo-pathologically. In the 13 cases of super paramagnetic iron peroxide, the sentinel node was identified each time. All biopsied lesions were removed, and in cancerous lesions, the surgical margins were healthy in all cases.

**Conclusion:** The MS localization technique is reliable and safe. For the patient, the main interest is a simplified procedure without long-term damage of the skin; for the radiologist, the rapidity of the procedure; for the surgeon, a real time guide for localizing the target; and for the hospital, an eased organization with regard to preoperative tracking during ambulatory surgery, with implantations possible up to 1 month prior to surgery, for instance at the time of the radiological review. The main limit to MS’s development remains its cost.

## INTRODUCTION

Thanks to widespread screening, the detection of non palpable lesions requiring surgery represents more than half of all breast surgeries, and the challenge of their surgical management lies in accurate preoperative location to allow complete removal, while limiting the sacrifice of healthy glandular tissue for a satisfactory aesthetic result^4^.

The reference preoperative location technique for the detection of non-palpable lesions of the breast is the radiologically placed hookwire. The effectiveness of this technique has long been established. It is reliable and inexpensive, but it has evolved little since the 20th century and has several drawbacks^7^. In particular, the insertion can be painful and a source of stress for the patient. Some placements cause haematoma or even, in rare cases, pneumothorax. The metal marker may migrate or be displaced before or during surgery. The presence of hematoma can alter the surgical removal and the anatomopathological result. In addition, the organization of its placement, the day before or the same day as the operation, may be logistically burdensome for the different services involved^4^. If the insertion is carried out the day before the operation, this implies that the patient goes home with the device and spends a night at home with the presence of this externalized wire, all of which may have an impact on the preoperative anxiety of the patients and her experience of the operation, as well as that of their family. In addition, the risk of displacement of the tip of the metallic marker may occur during the patient’s sleep, thus losing the benefit of the technique. Finally, the tip of the metallic marker which is in contact with the pectoral muscle can generate significant pain. For all these reasons, new techniques of preoperative marking have been developed. Magnetic seed (MS) appears to be a simple, non-radioactive and non-aggressive technique. MR takes the form of a 1x5mm paramagnetic clip made of iron oxide (Figure 1), visible on ultrasound and mammography and detected by a SentiMag^10^

**Figure 1.**
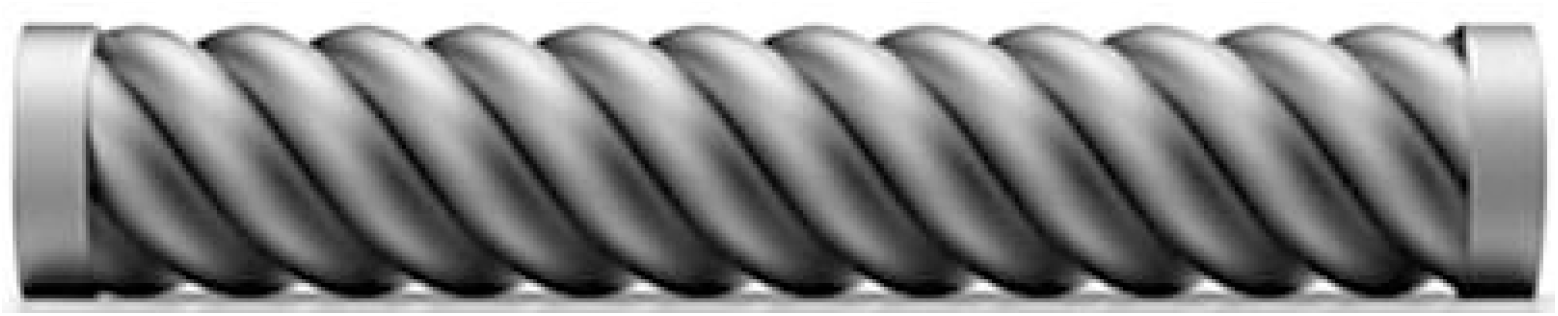
Magnetic seed (Magseed)

In 2018, ambulatory surgery represented 36% of all surgeries in France (the rates for 2019 and 2020 have not been raised due to the health situation caused by Covid 19). The objective of the HAS is to achieve a majority ambulatory practice of 70% in 2022^26^. The place of MR is particularly adapted in this new organization. The objective of this study is to evaluate the effectiveness and safety of magnetic seed (MS), performed under ultrasound, in the surgical management of non-palpable breast lesions in an essentially ambulatory surgical setting.

## MATERIALS AND METHODS

This study is a consecutive cohort performed at the Breast Center of Saint Joseph Hospital in Paris, France, between May 15 and December 21, 2018. Over this period, we studied the consecutive placement of 39 magnetic seed (MS) in 37 patients preoperatively, under ultrasound, within non-palpable breast lesions.Patients with benign, atypical or cancerous non-palpable lesions requiring surgical removal were included.

The placement of the MS was done under ultrasound control by an experienced radiologist (Figure 2). The tracer was inserted with a guide after local anesthesia. The magnetic tracer used in this study was approved for use in the localization of breast lesions up to 30 days before surgery ^11^.

**Figure 2.**
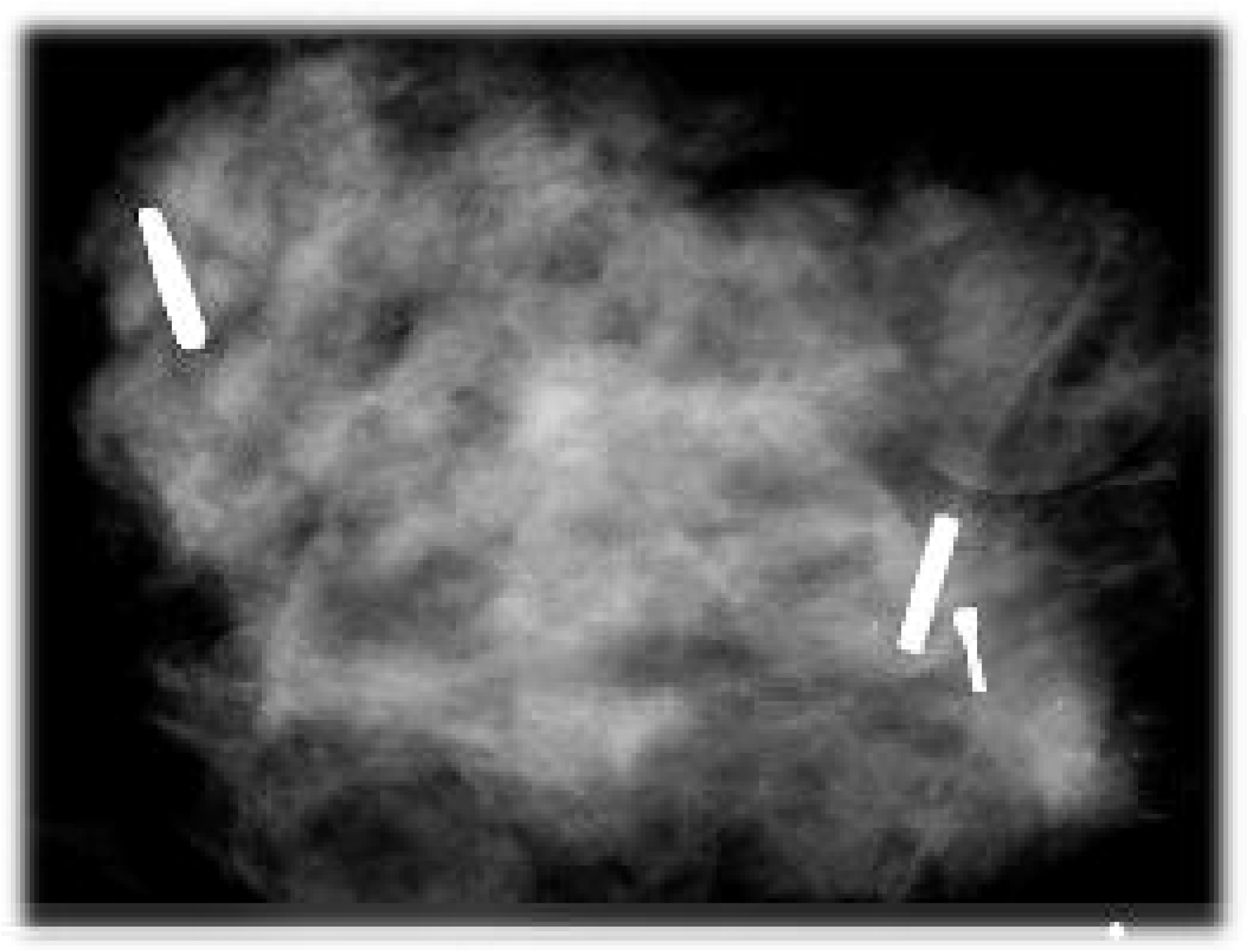
X-ray of the lesion with Magseed present (on the right)

When the tumor was an infiltrating ductal or lobular carcinoma, the tumor removal could be accompanied by a lymph node procedure; the realization of an axillary lymph node dissection or the dissection of the sentinel lymph node (SG). Two techniques were studied in this study for the dissection of the sentinel lymph node: patients over 60 years of age received an injection of iron peroxide (Sienna) and patients under 60 years of age or to be followed by breast MR received an isotope injection. In the operating room, the MR was located percutaneously with the magnetic detection probe. The ex-vivo magnetism was noted, and the surgical specimen was sent for radiography to confirm the presence of the MR and for traceability purposes (Figure 2).

The primary endpoint was complete removal of the lesion, with healthy margins. Secondary endpoints were the occurrence of complications, interference of MS with GS, and operative time from incision to specimen resection.

The ethics committee of Hospital Paris Saint-Joseph (Paris, France) gave ethical approval for this work.

## RESULTS

During the analysis period, 37 patients benefited from the MS technique. The average age of the patients was 57 years (33-86 years), the average size of the lesion on ultrasound was 12.7 mm (5-34 mm). All lesions were biopsied preoperatively. There were 29 infiltrating cancers, 7 atypical lesions, and 3 benign lesions. The time from landmark to surgery ranged from 0 to 21 days, with a mean of 5 days. Final lesion size averaged 12 mm, with lesions ranging from 3 to 29 mm. The mean time from incision to excision was 15 min for the first 10 cases. One patient with extensive papillomatous lesions had double MS placement. One patient had MS in each breast for bilateral invasive lobular carcinoma.

A lymph node procedure was required in 29 patients; the sentinel lymph node was identified by iron peroxide injection (Sienna) in 11 cases (45%), isotopes in 18 cases (55%) and axillary lymph node dissection was required in one patient. For the 11 cases in which the sentinel lymph node search was performed with Sienna+, the rate of identification of the GS was 100%, no interference with the use of MS was found.

The characteristics of the patients are summarized in Table 1.

**Table 1.** Characteristics of patients.

|  |  |
| --- | --- |
|  | (n = 37) |
| Age (year) | 57 (33-86ans) |
| Size on ultrasound (mm) | 12,8 (5-34mm) |
| Time to install magnetic tracking → surgery (days) | 0 to 21 |
| Incision - excision time (min) | 15 |
| Number of procedures associated with a sentinel node<br>Sienna + | 11 (28%) |
| Success | 39 (100%) |

The technical procedure for the location was characterized by a rapid placement, facilitated by an excellent brilliance of the needle and the marker. No pressure dressing was required, thus improving the performance and quality of the follow-up mammogram. All MS were placed within the target area during the follow-up mammogram.

For all patients studied, the MS was present in 38 out of 39 cases on room radiography. The only failure was due to the loss of the clip in the lumpectomy bed in a patient with a papillomatous tumor who was double located in the same breast. However, the target was well removed. All lesions were removed, with healthy margins in case of neoplastic lesion.

Of the 37 patients analyzed, we did not observe any postoperative complications.

## DISCUSSION

The study of this case series confirmed the efficacy and feasibility of the magnetic tracking technique. All lesions were removed with healthy margins. There were no postoperative complications. The logistics of this technique appear to be easier in practice than the metallic technique. We found a 100% efficiency on the use of 39 MS. In the light of our exploratory series, MS appears to be an effective and safe solution for preoperative location of non-palpable lesions of the breast and could thus be positioned as an alternative to hookwire localization.

The strength of this study is that there is currently no published study in France on the use, safety and efficacy of this modern method of detection in the surgical management of non-palpable breast lesions.

This is a single-center study with a limited number of patients, which limits the generalization of the results to other centers, but the implementation of innovative practices requires this type of study to define optimal management before allowing the diffusion of this technique. The evaluation of the efficiency (in terms of logistics, organizational ease) of MS was limited by the retrospective nature of our study, which inevitably led to some loss of data: the operating time was missing for the majority of patients, whereas some teams have shown that the use of new methods of identification made it possible to reduce operating time^25^. A prospective analysis, with a control group on the gold standard (metallic localization), would make it possible to evaluate the gain of time and the cost ratio of the MS technique.

Studies published in the literature since 2017 report efficacy of more than 90% in localizing subclinical breast lesions in cohorts ranging from 15 to 168 patients.

The safety of the technique had already been studied before its marketing in the United States. Thus, magnetic tracking received clearance in 2016 by the FDA after a feasibility study by Harvey & al^8^ who looked at 29 devices placed in 28 patients, 24 of which were under ultrasound and 5 under radiographic guidance. In this series, one patient had bilateral MR placement, 27 markers were placed directly on the target lesion and all implants were retrieved. But unlike our study, this study sought to study the migration of the landmarks and was performed on patients who were going to have a total mastectomy.

In 2019, in the team of Pohlodek et al. in Slovakia, 41 MS (Magseed) were placed in 38 patients. Twenty-seven patients with malignant tumors in this study had magnetic tracking simultaneously for tumor and sentinel node detection. All 38 breast lesions were accurately located using this method. No interference was observed on the magnetic probe between the tumor signals and the sentinel node tracer signals. All tumors were removed with healthy surgical margins^22^. The interaction between the two techniques could be evaluated in a larger series with the required statistical power.

The study by Reitsamer et al. showed in their study including 80 patients that the magnetic tracer is a reliable method for detecting post-chemotherapy target adenopathy. Nevertheless, one of the weaknesses of MS is that it cannot be used to detect cancerous lesions in the neoadjuvant setting because of signal artifacts on MRI, related to the presence of magnetic material in the breast. Also, other methods are being developed, such as the Savi Scout, an alternative tracking method that uses electromagnetic wave reflectors, but in a recent study showed difficulty in skin detection, inactivation of the tracer by contact with the electrocoagulation of the electric scalpel in the operating room, as well as migration of several of their tracers without identified cause^23^. Markers using radiofrequency are also being developed, which would also give the major advantage of not giving artifact to MRI, and would therefore allow the radiologist to place the marker at the same time as performing the diagnostic biopsy regardless of the stage and severity of the initial disease. New magnetic markers are being developed, including Sirius, which is distributed at a much lower price than Magseed with the same advantages and good results; studies are underway. On the other hand, although we have not noted any failure of the technique, we know that the MS may not emit a signal if it is placed at a distance greater than 4 cm from the target, which could pose a problem for large breasts. Finally, there is a technical difficulty: surgical steel instruments cannot be used because they produce an artifact on the Sentimag detection probe. For this reason, non-magnetic instruments are used, such as plastic or titanium retractors, which can generate additional costs^24^.

For the radiologist, MS has several advantages. First, because the placement can be done remotely from the procedure, the organization in the radiology department is simplified. Secondly, the time required for the dressing of the metal marker, often as long as the insertion of the device itself, is subtracted. Thirdly, the patient does not have to fast on the day of insertion (because she is at a distance from the surgical procedure) and anxiety is also less important. In 2021, Micha et al. compared the effectiveness and satisfaction of patients and physicians with metal and magnetic retrieval (a total of 296 patients). The results showed that patients experienced less anxiety at the time of tracking and at surgery in the magnetic tracking group, and physicians expressed greater ease of use of magnetic tracking. This is the first large study of satisfaction for localization for impalpable breast lesions.

One of the main disadvantages of MS is its cost, which is an obstacle to its deployment. While metal detection is a low-cost method (30 euros on average) and is fully covered by social security. The cost of magnetic identification is 410 euros (including VAT) and is borne by the establishment. On the other hand, the logistical simplification brought by this magnetic tracer could lead to savings and increase the satisfaction of the patient, as well as the medical team taking care of her. In addition, the price of MS is not fixed between different countries. A Dutch study^20^ studied the budgetary impact of MS according to its price in the management of a non-palpable breast tumor, the price ranged from €100 to €500, and the results show that if the price of MS does not exceed €175, it appears to be a cost-effective method by reducing the costs of organization, implementation and intervention. Manufacturers should take this aspect into account when determining and harmonizing the price of MS. Medico-economic studies (budgetary impact analysis comparing MR and metallic detection) and patient satisfaction studies (PROM’s Patient Reported Outcome measurements) are necessary to recommend the use of MR for preoperative detection of non-palpable breast lesions on a routine basis, and even to obtain its reimbursement.

## CONCLUSION

The technique of magnetic detection of subclinical breast lesions is reliable and safe. The major interest for the patient is an easy placement without any lasting invasion of the skin; for the radiologist: rapidity of the procedure; for the surgeon: real time guidance to find the target; and for the hospital: simplified organization of preoperative localization. With the development of outpatient care and minimally invasive surgery, this surgical innovation has its place by simplifying the organization, but its cost remains a barrier to its deployment

## Data Availability

Data cannot be shared publicly. Data are available from the Institutional Data Access / Ethics Committee (contact via corresponding author) for researchers who meet the criteria for access to confidential data.

